# Atrial Fibrillation and Risk of Injurious Falls Among Adults With Hypertension: A Secondary Analysis of SPRINT

**DOI:** 10.64898/2026.09.15.26363178

**Authors:** Tarek Zaho, Asem M. Mohsen, Moustafa Elnewishy, Mohamed Farag, Richard Kazibwe, Prashant D. Bhave, Elsayed Z. Soliman

## Abstract

**Background:** Atrial fibrillation (AF) may increase susceptibility to falls, but its association with injurious falls among adults with hypertension and whether intensive blood pressure (BP) lowering modifies this association are uncertain.

**Methods:** We analyzed 8,807 SPRINT participants with a baseline electrocardiogram and postrandomization follow-up for injurious falls. AF was evaluated at baseline and as a time-updated exposure incorporating AF identified during follow-up. Cox models estimated hazard ratios (HRs) and 95% CIs for the first injurious fall and assessed effect modification by randomized BP treatment assignment.

**Results:** Among 8,807 participants, 163 had baseline AF, 142 developed AF during follow-up, and 637 experienced an injurious fall. In fully adjusted analyses, baseline AF (HR, 1.79 [95% CI, 1.18–2.70]) and time-updated AF (HR, 1.89 [95% CI, 1.32–2.71]) were associated with higher fall risk. The association with time-updated AF was similar with standard and intensive BP treatment (HR, 1.91 [95% CI, 1.12–3.26] and 1.94 [95% CI, 1.18–3.19], respectively; P for interaction=0.964) and across prespecified subgroups. Results were robust in competing-risk and temporal-lag sensitivity analyses.

**Conclusions:** Among adults with hypertension and elevated cardiovascular risk, AF was independently associated with higher injurious-fall risk. The association was observed with baseline and time-updated AF and was not modified by intensive BP lowering, suggesting that AF identifies patients at increased fall risk without evidence that intensive BP treatment amplifies this risk.

## Introduction

Injurious falls are an important cause of morbidity among older adults and may complicate the management of hypertension.^1^ This issue is particularly relevant as recent guidelines have adopted lower blood pressure (BP) targets.² Identifying cardiovascular conditions associated with injurious falls may improve risk stratification and help inform BP management.

Atrial fibrillation (AF) is associated with reduced exercise capacity, impaired physical function, dizziness, cognitive impairment, and frailty, factors that may increase susceptibility to falls.³ However, the association between AF and incident injurious falls among adults with hypertension remains poorly defined. Previous studies have largely evaluated AF at baseline without accounting for AF developing during follow-up.^3^ Moreover, whether intensive BP lowering modifies the association between AF and injurious falls is unknown.

The Systolic Blood Pressure Intervention Trial (SPRINT) provides an opportunity to address these questions because it prospectively ascertained serious injurious falls, included serial electrocardiographic assessments allowing identification of baseline and incident AF, and randomized participants to intensive or standard systolic BP treatment.^6, 7^ These features permit evaluation of AF as a time-updated exposure and assessment of whether randomized BP treatment intensity modifies the association between AF and injurious falls.

Using data from SPRINT, we examined the associations of baseline and time-updated AF with the risk of a first injurious fall and assessed whether the association differed by randomized BP treatment assignment. We hypothesized that AF would be independently associated with a higher risk of injurious falls and that this association would be consistent across intensive and standard BP treatment groups.

## Methods

### Study Design and Population

This study was a secondary analysis of participants enrolled in the Systolic Blood Pressure Intervention Trial (SPRINT), a multicenter randomized clinical trial comparing intensive versus standard systolic blood pressure (BP) treatment. SPRINT enrolled adults aged ≥50 years with hypertension and increased cardiovascular risk but without diabetes mellitus or a history of stroke.^6,7^

Of the 9,361 randomized participants, 461 without a baseline electrocardiogram (ECG) were excluded because baseline AF status could not be ascertained. An additional 93 participants without postrandomization follow-up for injurious falls were excluded, resulting in a final analytic cohort of 8,807 participants (Figure 1). ECG quality codes were not used as an exclusion criterion. A history of falls before enrollment was not available as a standardized baseline variable in the released SPRINT data and therefore was not used as an exclusion criterion.

**Figure 1.**
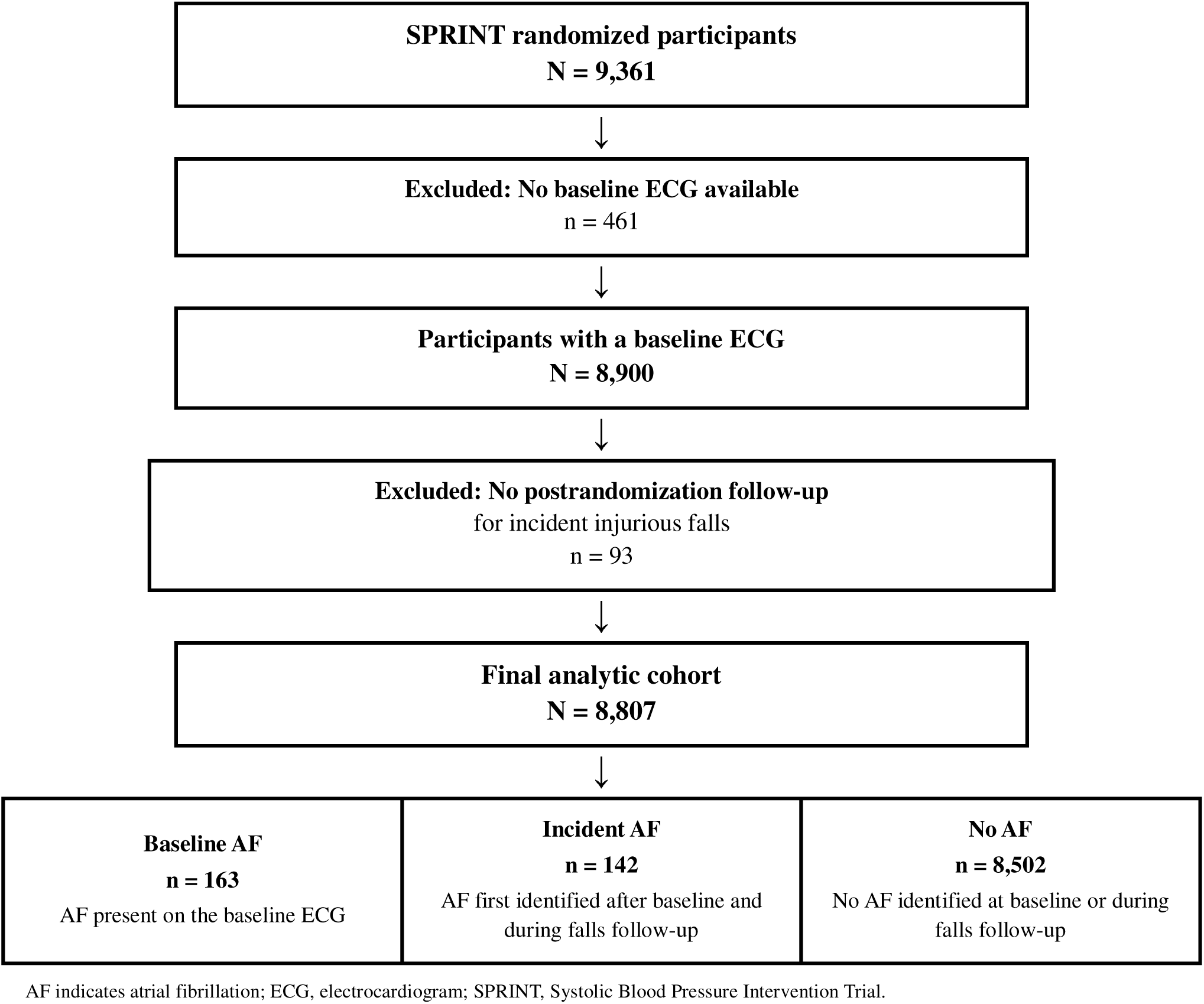
Selection of the Analytic Cohort and Classification of Atrial Fibrillation Status.

Participants were followed from randomization until the first validated injurious fall, death, last known follow-up, or the end of study follow-up, whichever occurred first.

### Ascertainment of Atrial Fibrillation

AF was ascertained from protocol 12-lead ECGs obtained at baseline, year 2, year 4, and the closeout visit. ECGs were digitally recorded using GE MAC 1200 electrocardiographs (GE Healthcare) at standard calibration (10 mm/mV and 25 mm/s) and transmitted to the Epidemiological Cardiology Research Center (EPICARE) at Wake Forest University School of Medicine for centralized processing.

ECG quality was reviewed by trained technicians blinded to randomized treatment assignment before automated processing using the GE Marquette 12-SL algorithm (version 2001). ECG abnormalities were classified according to the Minnesota Code.^8^ ECGs classified as AF were visually confirmed by trained ECG readers according to previously described SPRINT procedures.^8^ For participants with AF identified on more than one ECG, the earliest occurrence was used to define AF onset.

Two complementary AF exposure definitions were examined. Baseline AF was defined as AF identified on the baseline ECG and was modeled as a time-fixed exposure. Participants were classified according to their baseline AF status throughout follow-up, regardless of AF subsequently identified on follow-up ECGs.

Time-updated AF incorporated both baseline and incident AF. Participants with AF at baseline were considered exposed from randomization. Participants without baseline AF contributed AF-unexposed person-time until AF was first identified on a follow-up protocol ECG and AF-exposed person-time thereafter.^1^ Participants in whom AF was not identified on any available protocol ECG remained unexposed throughout follow-up.

For the time-updated analysis, follow-up was structured in counting-process start–stop format, allowing AF exposure status to change at the time AF was first identified during follow-up.

### Ascertainment of Injurious Falls

The primary outcome was the first incident injurious fall after randomization, ascertained from prospectively collected SPRINT safety-event data using the validated injurious-fall indicator. Only falls occurring after randomization were included. Among participants with multiple injurious falls, the first event was used for the primary time-to-event analysis.

### Covariates

Covariates were selected a priori based on their established associations with AF, injurious falls, cardiovascular risk, or BP treatment and their potential to confound the association of interest. These included age, sex, race, randomized BP treatment assignment, educational attainment, smoking status, alcohol consumption, physical activity, body mass index, baseline systolic BP, number of antihypertensive medications, total cholesterol to high-density lipoprotein cholesterol ratio, triglyceride level, estimated glomerular filtration rate (eGFR), urine albumin-to-creatinine ratio (UACR), and prevalent cardiovascular disease. UACR was natural log-transformed because of its right-skewed distribution.^11^ Variables considered potential consequences of AF or intermediates in the association between AF and injurious falls were not included in the primary multivariable model.

### Statistical Analysis

Baseline characteristics were summarized according to 3 mutually exclusive AF categories: no AF during follow-up, baseline AF, and incident AF identified during follow-up. Participants with baseline AF were excluded from the incident-AF group, and incident AF required absence of AF at baseline. Continuous variables were presented as mean (SD) or median (IQR), as appropriate, and categorical variables as number (percentage). Differences across groups were assessed using analysis of variance or the Kruskal-Wallis test for continuous variables and the χ² or Fisher exact test for categorical variables, as appropriate.

Cox proportional hazards regression was used to estimate hazard ratios (HRs) and 95% CIs for the first incident injurious fall, with time since randomization as the underlying time scale. Conventional Cox models were fitted for baseline AF. For time-updated AF, extended Cox models were fitted using counting-process start–stop data, allowing AF exposure status to change when AF was first identified during follow-up.

Three sequential models were examined for each AF exposure definition. Model 1 was unadjusted. Model 2 adjusted for age, sex, race, and randomized BP treatment assignment. Model 3 additionally adjusted for educational attainment, smoking status, alcohol consumption, physical activity, body mass index, baseline systolic BP, number of antihypertensive medications, total cholesterol to high-density lipoprotein cholesterol ratio, triglyceride level, estimated glomerular filtration rate, log-transformed urine albumin-to-creatinine ratio, and prevalent cardiovascular disease. Each model included participants with complete data for the covariates included in that model; therefore, sample sizes and event counts are reported separately for each model. The proportional hazards assumption was evaluated using time-by-exposure interaction terms and graphical assessment of scaled Schoenfeld residuals, as appropriate. Potential influential observations were assessed using score and deviance residuals.

The association between time-updated AF and injurious falls was examined within prespecified subgroups defined by age (<75 versus ≥75 years), sex, race (Black versus non-Black), baseline systolic BP (<140 versus ≥140 mm Hg), randomized BP treatment assignment (intensive versus standard), chronic kidney disease (estimated glomerular filtration rate <60 versus ≥60 mL/min/1.73 m²), prevalent cardiovascular disease, and obesity (body mass index ≥30 versus <30 kg/m²). Fully adjusted time-updated Cox models were fitted within each subgroup. Effect modification was assessed by including multiplicative interaction terms between time-updated AF and each subgroup variable in the fully adjusted model, with interaction P values obtained from Wald tests. The variable defining each subgroup was omitted from the corresponding stratified model.

Simon–Makuch methods were used to estimate the cumulative probability of a first injurious fall according to current AF status. Participants without AF at baseline initially contributed to the AF-free group and switched to the AF group when AF was first identified during follow-up. Because conventional log-rank testing is not appropriate for a time-updated exposure, statistical inference was based on the time-updated Cox model.

As a sensitivity analysis, 30– and 90-day lags were introduced after incident AF identification, such that participants remained classified as AF-unexposed until 30 or 90 days after AF was first identified, to reduce the possibility that AF detection was temporally related to the evaluation of a fall or another acute event. Also, participants with baseline AF were excluded to examine the association of incident AF alone with injurious falls. Finally, Fine–Gray subdistribution hazards regression was used to evaluate the association between baseline AF and injurious falls while treating death as a competing event.

All statistical tests were 2-sided, with P<0.05 considered statistically significant. Analyses were performed using SAS 9.4 [ SAS Institute, Cary, NC]

## Results

The analytic cohort included 8,807 SPRINT participants with a baseline ECG and postrandomization follow-up for injurious falls (Figure 1). Of these, 163 (1.9%) had AF at baseline, 142 (1.6%) developed AF during follow-up, and 8,502 (96.5%) had no AF identified during observed follow-up. During a median follow-up of 3.75 years (IQR, 3.21–4.28), 637 participants experienced a first injurious fall, corresponding to an incidence rate of 20.4 per 1,000 person-years. Baseline characteristics according to AF status are presented in Table 1. Participants with baseline or incident AF were older and generally had a less favorable cardiovascular risk profile than participants without AF.

**Table 1.** Baseline Characteristics According to Atrial Fibrillation Status.

| Characteristic | No AF during follow-up (n=8,502) | Baseline AF (n=163) | Incident AF during follow-up (n=142) | P value |
| --- | --- | --- | --- | --- |
| Age, y | 67.6±9.3 | 75.8±8.3 | 72.9±9.2 | <0.001 |
| Age ≥75 y | 2,304 (27.1) | 101 (62.0) | 69 (48.6) | <0.001 |
| Women | 3,049 (35.9) | 31 (19.0) | 34 (23.9) | <0.001 |
| Black race | 2,742 (32.3) | 15 (9.2) | 20 (14.1) | <0.001 |
| Intensive BP treatment | 4,259 (50.1) | 96 (58.9) | 58 (40.8) | 0.007 |
| Baseline systolic BP, mm Hg | 145.1±11.2 | 143.4±10.4 | 144.8±11.8 | 0.138 |
| Body mass index, kg/m<sup>2</sup> | 29.8±5.6 | 29.8±5.2 | 30.0±5.8 | 0.892 |
| Current smoking | 1,222 (14.4) | 6 (3.7) | 11 (7.7) | <0.001 |
| Alcohol use | 5,460 (64.3) | 100 (61.3) | 107 (75.4) | 0.017 |
| Education level | 7.7±2.4 | 8.0±2.6 | 7.9±2.5 | 0.243 |
| Physical activity score | 2.9±1.4 | 2.7±1.5 | 2.8±1.5 | 0.192 |
| BP medications (n) | 1.8±1.0 | 2.2±1.0 | 2.2±1.0 | <0.001 |
| Cholesterol/HDL ratio | 3.8±1.2 | 3.5±0.9 | 3.6±1.0 | <0.001 |
| Triglycerides, mg/dL | 107.0 (77.0–150.0) | 95.0 (71.5–135.5) | 98.5 (74.2–135.0) | 0.010 |
| eGFR, mL/min/1.73 m<sup>2</sup> | 72.1±20.6 | 63.6±18.9 | 67.7±17.1 | <0.001 |
| Urine albumin-creatinine ratio, mg/g | 9.4 (5.6–20.6) | 26.3 (10.8–63.6) | 11.9 (7.0–27.6) | <0.001 |
| Chronic kidney disease | 2,347 (27.7) | 77 (47.2) | 49 (34.5) | <0.001 |
| Cardiovascular disease | 1,637 (19.3) | 64 (39.3) | 49 (34.5) | <0.001 |

Values are mean±SD, median (interquartile range), or n (%). AF indicates atrial fibrillation; BP, blood pressure; eGFR, estimated glomerular filtration rate; and HDL, high-density lipoprotein. P values compare the 3 AF groups and are descriptive.

Both baseline and time-updated AF were associated with a higher risk of a first injurious fall (Table 2). In unadjusted analyses, baseline AF was associated with an HR of 2.75 (95% CI, 1.86–4.08), which was attenuated but remained significant after full multivariable adjustment (HR, 1.79; 95% CI, 1.18–2.70; P=.006). Similarly, time-updated AF was associated with an HR of 2.86 (95% CI, 2.02–4.04) in the unadjusted model and an HR of 1.89 (95% CI, 1.32–2.71; P<.001) after full adjustment **(Table 2).**

**Table 2.** Association of Atrial Fibrillation With First Incident Injurious Fall.

| AF exposure | Person-years | Injurious falls, n | Incidence rate per 1,000 PY | HR (95% CI) | Pvalue |
| --- | --- | --- | --- | --- | --- |
| **Baseline AF** |  |  |  |  |  |
| No AF | 30,687 | 611 | 19.9 | 1.00 (Reference) | — |
| AF | 475 | 26 | 54.8 |  |  |
| Model 1 |  |  |  | 2.75 (1.86–4.08) | <0.001 |
| Model 2 |  |  |  | 1.93 (1.30–2.88) | 0.001 |
| Model 3 |  |  |  | 1.79 (1.18–2.70) | 0.006 |
| **Time-updated AF** |  |  |  |  |  |
| AF-unexposed person-time | 30,559 | 603 | 19.7 | 1.00 (Reference) | — |
| AF-exposed person-time | 603 | 34 | 56.3 |  |  |
| Model 1 |  |  |  | 2.86 (2.02–4.04) | <0.001 |
| Model 2 |  |  |  | 2.01 (1.42–2.86) | <0.001 |
| Model 3 |  |  |  | 1.89 (1.32–2.71) | <0.001 |

Incidence rates are crude and expressed per 1,000 person-years. For the time-updated analysis, participants without AF at baseline contributed AF-unexposed person-time until AF was first identified during follow-up and AF-exposed person-time thereafter. Model 1 is unadjusted. Model 2 adjusts for age, sex, race, and randomized BP treatment assignment. Model 3 additionally adjusts for educational attainment, smoking status, alcohol consumption, physical activity, body mass index, baseline systolic BP, number of antihypertensive medications, total cholesterol to high-density lipoprotein cholesterol ratio, triglycerides, estimated glomerular filtration rate, log-transformed urine albumin-to-creatinine ratio, and prevalent cardiovascular disease.

Simon–Makuch estimates demonstrated a higher cumulative probability of a first injurious fall during AF-exposed than AF-free person-time **(Figure 2),** consistent with the association observed in the fully adjusted time-updated Cox model.

**Figure 2:**
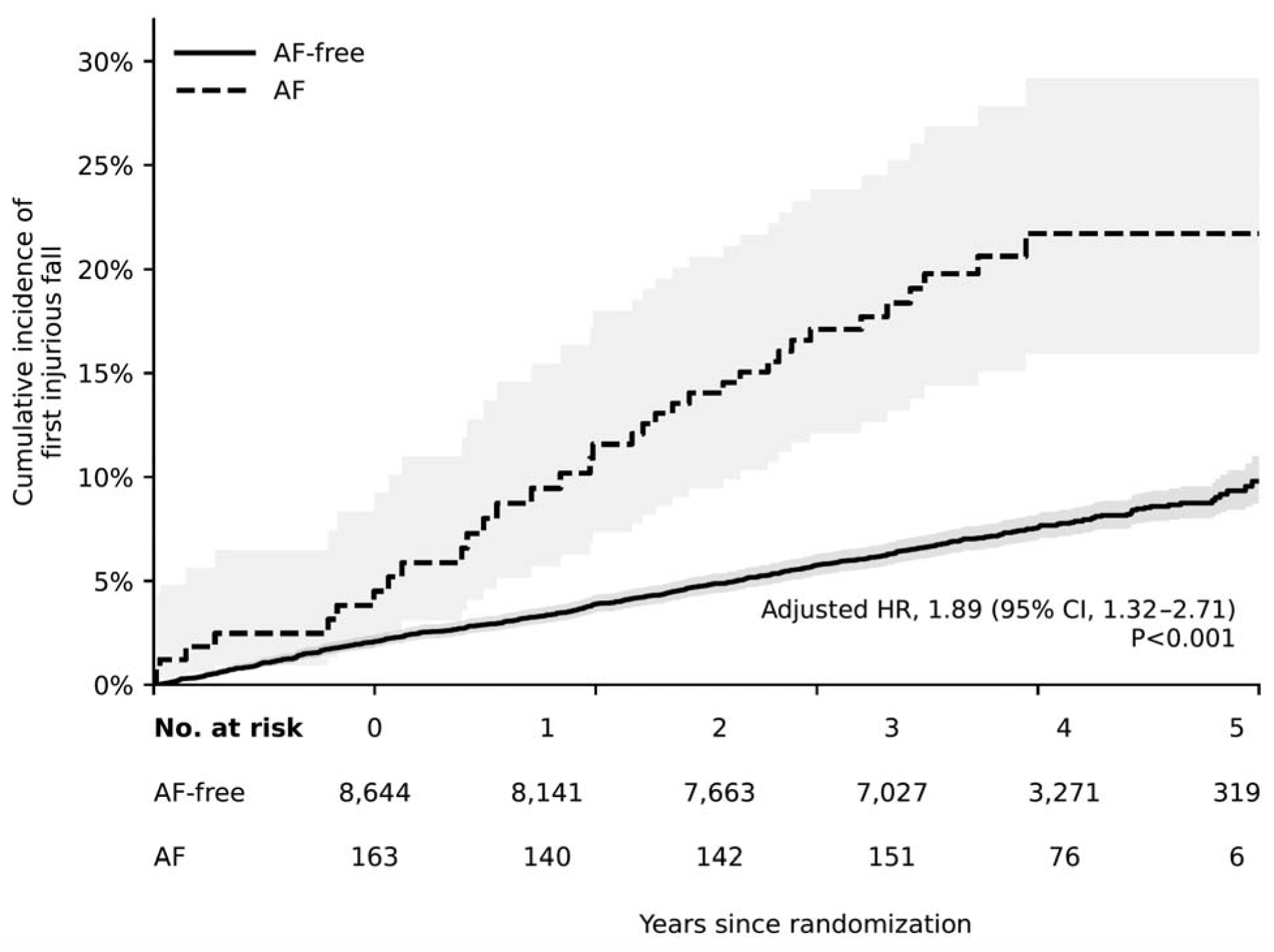
Cumulative Incidence of First Injurious Falls According to Time-Updated Atrial Fibrillation Status. Simon–Makuch estimates are shown according to current AF status. Participants without baseline AF contributed AF-free person-time until first AF detection and AF-exposed person-time thereafter; participants with baseline AF were AF-exposed from randomization. Numbers at risk are shown at yearly intervals. Statistical inference is based on the fully adjusted time-updated Cox model.

The association between time-updated AF and injurious falls was generally consistent across prespecified subgroups defined by age, sex, race, baseline systolic BP, randomized treatment assignment, chronic kidney disease, prevalent cardiovascular disease, and obesity, with no evidence of effect modification (all P for interaction >.05) **(Figure 3)**. Of particular relevance to the primary study question, the association was similar among participants randomized to standard BP treatment (HR, 1.91; 95% CI, 1.12–3.26) and intensive BP treatment (HR, 1.94; 95% CI, 1.18–3.19; P for interaction=.964).

**Figure 3:**
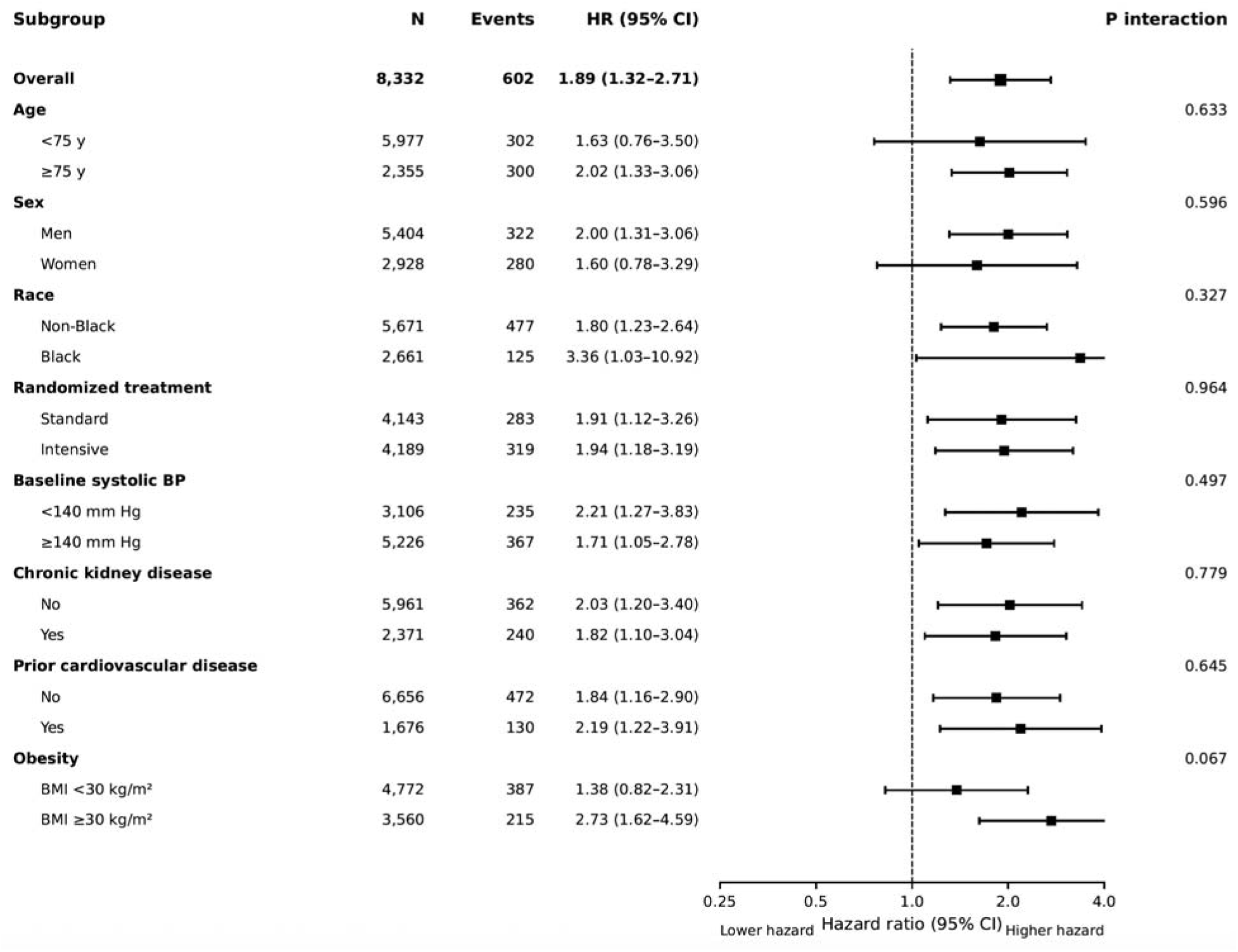
Association of Time-Updated Atrial Fibrillation With First Incident Injurious Fall Across Prespecified Subgroups. Squares represent hazard ratios and horizontal lines represent 95% CIs. Estimates are from adjusted time-updated Cox models. The variable defining each subgroup was omitted from the corresponding stratified model. AF indicates atrial fibrillation; BP, blood pressure; BMI, body mass index.

Results were robust to sensitivity analyses. When AF exposure was delayed by 30 and 90 days after incident AF identification, the fully adjusted HRs were 1.91 (95% CI, 1.34–2.73; P<.001) and 1.90 (95% CI, 1.32–2.72; P<.001), respectively. After excluding participants with AF at baseline, incident AF remained associated with a higher risk of subsequent injurious falls (HR, 2.20 [95% CI, 1.09–4.47]; P=.029), although only 8 injurious falls occurred after incident AF onset. In a Fine–Gray competing-risk analysis treating death as a competing event, the association between baseline AF and injurious falls was essentially unchanged (subdistribution HR, 1.78 [95% CI, 1.18–2.68]; P=.006).

## Discussion

In this analysis of adults with hypertension enrolled in SPRINT, AF was independently associated with a higher risk of injurious falls. After adjustment for demographic characteristics, lifestyle factors, and cardiovascular risk factors, baseline AF was associated with a 79% higher risk of injurious falls, while time-updated AF was associated with an 89% higher risk. The findings were robust after accounting for competing mortality, introducing temporal lags after incident AF identification, and restricting the analysis to incident AF. Importantly, the association between AF and injurious falls did not differ between participants randomized to intensive versus standard BP treatment. These findings suggest that AF identifies a subgroup of adults with hypertension at increased risk of injurious falls, without evidence that intensive BP lowering amplifies this risk.

The relationship between AF and injurious falls has been less extensively studied than its established associations with stroke, heart failure, and mortality. Previous studies have linked AF with impaired physical function, reduced exercise tolerance, frailty, and cognitive decline,^4, 13^ but direct evidence regarding injurious falls remains limited. Moreover, studies based solely on baseline AF status do not account for AF developing during follow-up. In the present study, the association was observed using both baseline and time-updated AF, with somewhat stronger estimates when AF status was updated during follow-up. Importantly, after participants with baseline AF were excluded, incident AF remained associated with subsequent injurious falls, supporting an association that was not driven solely by individuals with AF already present at study entry.

Several mechanisms may contribute to the association between AF and injurious falls. Loss of atrial contribution to ventricular filling and an irregular or rapid ventricular response may reduce cardiac output and contribute to fatigue, dizziness, or transient cerebral hypoperfusion.^12, 13^ AF also commonly coexists with frailty, sarcopenia, and reduced physical activity, which may adversely affect gait and postural stability.^14,15^ Cognitive impairment and cerebral microvascular injury associated with AF may further impair executive function and the ability to respond to environmental hazards.^16,17^ Medication burden, including rate– or rhythm-control therapies that may contribute to bradycardia or hypotension, represents another potential pathway.^18^ These mechanisms are not mutually exclusive, and AF may represent both a contributor to falls and a marker of broader cardiovascular and functional vulnerability.

An important objective of this study was to determine whether intensive BP lowering modified the association between AF and injurious falls. Concerns about hypotension, medication burden, and reduced cerebral perfusion could raise the possibility that patients with AF are particularly susceptible to falls during intensive BP treatment. However, the associations were nearly identical among participants randomized to standard and intensive treatment, with no evidence of interaction. These findings suggest that the excess risk of injurious falls associated with AF does not appear to be amplified by intensive BP lowering. Importantly, this analysis addresses whether AF modifies susceptibility to injurious falls according to randomized BP treatment intensity, rather than the overall effect of intensive BP lowering on fall risk, which has been addressed in previous SPRINT analyses.^19^

Several sensitivity analyses supported the robustness of the findings.^20,21^ In the Fine– Gray analysis treating death as a competing event, the association between baseline AF and injurious falls was essentially unchanged, suggesting that differential mortality was unlikely to explain the association. Introducing 30– and 90-day lags after incident AF identification also had little effect on the results, reducing concern that AF was identified during evaluation of a fall or a temporally related acute event. Furthermore, after excluding participants with AF at baseline, incident AF remained associated with a higher risk of subsequent injurious falls (HR, 2.20 [95% CI, 1.09–4.47]). This latter estimate should nevertheless be interpreted cautiously because only 8 injurious falls occurred after incident AF onset, resulting in limited precision.

These findings may have clinical implications. AF is routinely identified in cardiovascular practice and may provide readily available information relevant to fall-risk assessment among adults with hypertension.^22^ The consistency of the association across prespecified subgroups suggests that it is not confined to a particular demographic or cardiovascular risk subgroup. Patients with AF, particularly those with additional indicators of frailty, impaired mobility, or orthostatic symptoms, may warrant greater attention to fall risk, including assessment of gait, balance, and medication burden. At the same time, the absence of interaction with randomized BP treatment assignment provides reassurance that the association between AF and injurious falls was not greater among participants receiving intensive BP treatment. Whether incorporating AF into established fall-risk assessment approaches improves risk prediction or whether targeted fall-prevention interventions benefit patients with AF warrants further investigation.

### Strengths and Limitations

This study has several strengths. SPRINT provided a large, well-characterized cohort of adults with hypertension, standardized prospective follow-up and prospective ascertainment of serious injurious falls. AF was ascertained from serial protocol ECGs with centralized processing and confirmation, allowing AF to be evaluated both at baseline and as a time-updated exposure. Randomized assignment to intensive versus standard BP treatment provided a unique setting to evaluate whether BP treatment intensity modified the association between AF and injurious falls. The consistency of the findings across prespecified subgroups, competing-risk analyses, temporal-lag analyses, and the analysis restricted to incident AF further supports the robustness of the findings.

Several limitations should also be acknowledged. Although BP treatment assignment was randomized, AF was not; therefore, the association between AF and injurious falls is observational and cannot establish causality, and residual or unmeasured confounding remains possible. SPRINT did not systematically collect several potentially relevant factors, including standardized baseline frailty measures, formal gait or balance assessments, and a validated history of falls before enrollment. AF ascertainment was based on protocol ECGs obtained at discrete time points rather than continuous rhythm monitoring; therefore, paroxysmal or asymptomatic AF occurring between assessments may have gone undetected, and the time of first ECG identification may not correspond precisely to AF onset. Injurious falls were ascertained as safety events rather than through a dedicated falls-surveillance protocol, potentially resulting in underascertainment of less severe events. Although the incident-AF-only analysis supported the primary findings, only 8 injurious falls occurred after incident AF onset, limiting precision. Finally, SPRINT enrolled adults with hypertension and elevated cardiovascular risk and excluded individuals with diabetes mellitus, prior stroke, and recent symptomatic heart failure, which may limit generalizability to these and lower-risk populations.

### Conclusions

Among adults with hypertension and elevated cardiovascular risk enrolled in SPRINT, AF was independently associated with a higher risk of injurious falls. The association was observed with both baseline and time-updated AF, remained present when the analysis was restricted to incident AF, and did not differ according to randomized BP treatment assignment. These findings suggest that AF may identify hypertensive adults at increased risk of injurious falls, while providing no evidence that intensive BP lowering further increases AF-associated fall risk.

## Data Availability

All data produced are available online at SPRINT

## Acknowledgements

Data for this study were obtained from the publicly available SPRINT data repository through the National Heart, Lung, and Blood Institute (NHLBI) Biologic Specimen and Data Repository Information Coordinating Center (BioLINCC). The present analysis was conducted independently of the SPRINT Research Group, and the conclusions reported herein do not necessarily represent the views of the SPRINT investigators or the NHLBI.

## Competing interest

Authors declare no related competing interests.

## Availability of data and materials

The SPRINT data used in this analysis are publicly available through the National Heart, Lung, and Blood Institute (NHLBI) Biologic Specimen and Data Repository Information Coordinating Center (BioLINCC) at https://biolincc.nhlbi.nih

## Notes

### Competing Interest Statement

The authors have declared no competing interest.

